# Understanding reported COVID-19 cases in England following changes to testing, between November 2021 and April 2022

**DOI:** 10.1101/2022.06.28.22276549

**Authors:** Florence Halford, Sophie Nash, Elise Tessier, Meaghan Kall, Gavin Dabrera

**Affiliations:** UK Health Security Agency

## Abstract

**Background:** Over the course of the pandemic, testing policies for SARS-CoV-2 have varied considerably in England, particularly in the five months up to 1 April 2022 when free community testing ended. We described the trends and demographics of COVID-19 cases during this period.

**Methods:** COVID-19 cases reported between 15 November 2021 and 30 April 2022 were extracted and aggregated by testing pillar: Pillar 1 for those tested within the NHS, private or public health laboratories, and Pillar 2 for community testing. COVID-19 cases were described by epi-week, and stratified by test type, age, sex, index of multiple deprivation (IMD), region, and population density. Incidence rates were also calculated and stratified by IMD and region.

**Results:** Of 10,196,425 COVID-19 cases, 7.3% were reported under Pillar 1 and 92.7% under Pillar 2. From 15 November 2021 to 31 March 2022, most Pillar 2 cases were tested either by polymerase chain reaction (PCR) only or PCR with lateral flow device (LFD) (70.8%) and three in ten cases tested using LFD only. However, between 1 April and 30 April 2022 this rose to nine out of ten cases testing using LFD only. Over the whole period studied and under both pillars, the majority of cases were female (55.2%), resided in the South East (17.0%) and in the age group 30-39 years (18.6%). Trends in IMD and population density varied over the period. When stratifying by IMD the highest case numbers and incidence rates reported under Pillar 1 and NHS were in those in the most deprived quintile. This was also seen for cases reported under Pillar 2 by LFD until 11 January 2022, where a reverse in the trend occurred with the highest cases and rates in the least deprived quintile. This same pattern was observed when describing the cases by population density, with Pillar 2 LFD reported cases being highest in the most densely populated regions until 11 January, from when there was a switch to the highest cases being in the least densely populated regions.

**Conclusion:** Differences and trends were observed in reported COVID-19 cases in England, particularly those tested under Pillar 2 following the introduction of testing policy changes. To better understand the impact of these changes over the course of the COVID-19 pandemic, as well as to predict the impact of future testing policies, it would be beneficial to investigate the accessibility of testing amongst different populations. Currently, Pillar 1 COVID-19 cases are likely to be more representative of symptomatic cases requiring testing for a clinical need, as these are less impacted by variations in testing patterns compared to Pillar 2. However, a limitation of that approach is that use of Pillar 1 alone would be biased towards those more likely to be clinically unwell.

## Introduction

In May 2020 the UK government introduced free of charge SARS-CoV-2 polymerase chain reaction (PCR) tests, initially only for symptomatic individuals, and then later expanded to being widely available for the general public.[1]

As the COVID-19 pandemic has evolved, so have testing recommendations. During this time, there have been two main “pillars” of testing: Pillar 1 focused mainly on testing for a clinical or public health need and was undertaken in NHS and public health laboratories, while being predominantly based on PCR testing.

A second pillar (Pillar 2) delivered mass community testing outlined above for symptomatic individuals; this included free laboratory based PCR testing and access to lateral flow device (LFD) kits which members of the public could access to undertake in their own home. Pillar 2 LFD results relied on the reporting of results by the users to a government web portal, while PCR tests from both pillars were reported by laboratories directly to public health agencies via notifiable disease regulations.

The UK government Living with COVID strategy [2, 3] outlined that on 1 April 2022 access to free lateral flow device (LFD) tests in England would be removed, while testing among symptomatic individuals has remained available in particular settings such as in National Health Service (NHS) hospitals, for NHS staff, and among those eligible for COVID-19 treatments.

Prior to April 2022, a number of step-by-step policy changes were made to allow for a gradual phase out of widespread free SARS-CoV-2 testing. On 5 January 2022 it was announced that PCR confirmatory tests for those who tested positive by LFD would end, with this coming into action on the 11 January [4]. At the time, it was still a legal requirement to self-isolate following a positive LFD result, due to the high specificity of LFD tests.[5] The following month it was also announced that from 24 February the legal requirement to isolate, following either a positive SARS-CoV-2 test or if an individual was a known contact of a positive case, would end. In addition, the self-isolation support payments would end along with the announcement that Statutory Sick Pay due to COVID-19 would be phased out with the remaining 1 April policy changes.[2]

The guidance for testing in populations at high risk of severe COVID-19 disease who are eligible for treatment, along with those who are inpatients or working in hospitals and adult social care settings, has remained the same over the time period of interest where free testing continues to be available for these populations.

We investigated trends in testing from 15 November 2021 to 30 April 2022, with a focus on the different test types used and between key dates when policy decisions around testing were made. The aim of this study is to understand the socio-demographic characteristics of people who have tested positive for SARS-CoV-2 during this period.

## Methods

Confirmed COVID-19 cases from 15 November 2021 to 30 April 2022 were extracted on 3 May 2022. Key dates of interest where policy changes to testing occurred were selected as ‘cut off points’, where changes in case numbers were expected.

Testing in England has been aggregated by “pillars” where tests coming from Pillar 1 are privately purchased tests, or NHS hospital admissions and occupational screening of healthcare workers, and tests coming from Pillar 2 are PCR and LFD tests within the community.

COVID-19 cases by epi-week and pillar test type were described by each of the following covariates: age group, sex, index of multiple deprivation (IMD) quintiles from 2019[6], regions, population density estimates from 2019[7], and levels of urbanisation. Trends in the total number of cases per 100,000 population were assessed using Office for National Statistics (ONS) 2020 population estimates.[8]

Age standardised incidence rates were calculated to adjust for differences in age structures and using population level data to calculate incidence per 100,000 population. Age standardised mortality rates were also calculated using the same methods among COVID-19 cases who have died within 28 days of the first specimen date of the most recent infection or 28 days after the first specimen date and with COVID-19 mentioned on the death certificate, as described in the UKHSA mortality reports.[9] All analyses were conducted using STATA 17.[10]

## Results

A total of 10,196,425 COVID-19 cases reported to national surveillance systems with a specimen date from 15 November 2021 to 30 April 2022 were identified. Of which, 744,703 (7.30%) were in Pillar 1 and 9,451,722 (92.70%) were in Pillar 2 (Table 1).

**Table 1:** Number of COVID-19 cases by Pillar and test type between 15 November 2021 to 30 March 2022, 1 April 2022 – 30 April 2022, and overall from 15 November 2021 to 30 April 2022, England.

|  | Pillar 1 |  |  |  | Pillar 2 |  |  |
| --- | --- | --- | --- | --- | --- | --- | --- |
|  | *Private* | *NHS* | *Unknown type* | *Total Pillar 1* | *PCR, and PCR with LFD* | *LFD* | *Total Pillar 2* |
| **Case numbers (%)**<br>**15 November 2021 – 31 March 2022**<br>**(n=9,539,923)** | 285,035<br>(43.85) | 361,856<br>(55.66) | 3,204<br>(0.49) | 650,095<br>(100) | 6,296,895<br>(70.83) | 2,592,933<br>(29.17) | 8,889,828<br>(100) |
| **Case numbers (%)**<br>**1 April – 30 April 2022 (n=656,502)** | 27,772<br>(29.35) | 66,287<br>(70.06) | 549<br>(0.58) | 94,608<br>(100) | 57,009<br>(10.15) | 504,885<br>(89.85) | 561,894<br>(100) |

Since 1 April 2022, the number of cases reported through Pillar 2 dropped with a total of 561,894 cases (85.60% of total cases in April) between 1 April to 30 April 2022 (Table 1). The proportion of reported Pillar 2 cases which were detected by PCR and combined LFD/PCR decreased from 70.83% between 15 November 2022 to 31 March 2022 to 10.15% during the month of April 2022, whereas the proportion of LFD testing increased from 29.17% to 89.85% (Table 1).

### Positive cases by age

Overall, trends have varied among each age group between 15 November 2021 and 30 April 2022.

Among those detected under Pillar 1 NHS, the highest case numbers were seen in those of working age up to the end of January 2022, after which those over 80 years saw the highest case numbers (Figure 1A). The lowest case numbers tested under Pillar 1 NHS were seen in children (0-19 years), with the total weekly cases not exceeding 2,000 during the time period of interest.

**Figure 1:**
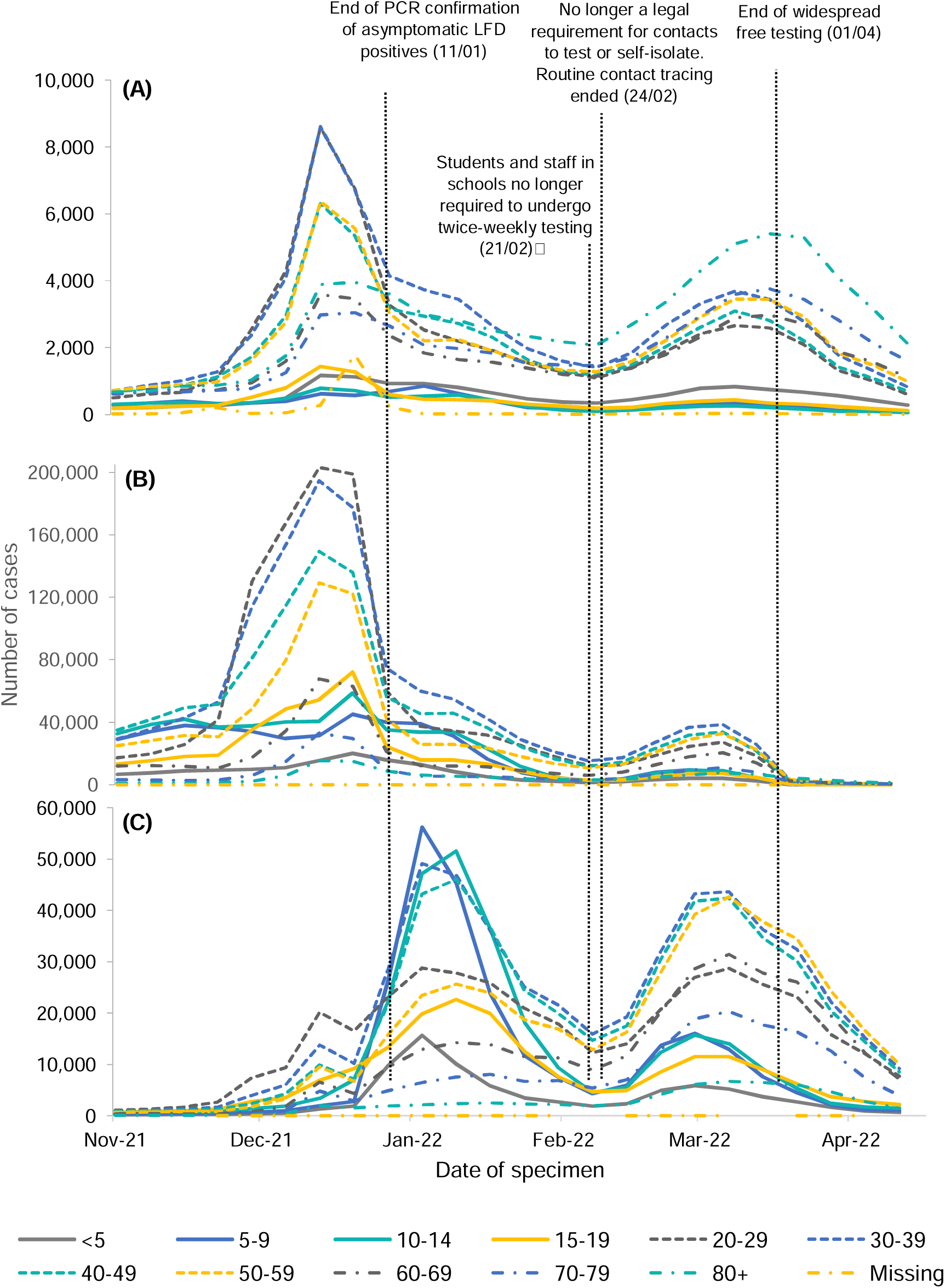
**Number of weekly COVID-19 cases* tested under (A) Pillar 1 NHS, (B) Pillar 2 PCR and PCR with LFD, and (C) Pillar 2 LFD only, by age group: 15 November 2021 to 30 April 2022, England.** *Dates of interest due to changes in testing policy are labelled*. *\* Please note Figures 1A-C display crude case numbers without taking into account population denominators*.

Figure 1B highlights a steep drop in cases detected through Pillar 2 PCR testing in January 2022, which coincides with an equally steep increase in Pillar 2 LFD (Figure 1C) cases. As shown in the figure, this corresponds with the end to PCR confirmatory testing of asymptomatic LFD positives on 11 January 2022.

Trends in both PCR and LFD tested cases began to decline among all age groups in the beginning of February 2022. As with positive cases reported through Pillar 1 NHS, the highest case numbers detected via Pillar 2 PCR and PCR with LFDs were also seen in those of working age.

From 21 February 2022, there was an increase in cases of all age groups and test types. This was followed by a decline in all age groups just before 1 April 2022, when the end to widespread free testing was introduced.

### Sex

Under all pillar breakdowns of cases by sex (Figure 2) females saw higher case numbers than males, except for those tested privately under Pillar 1 (Figure 2A). This difference was most apparent in those detected via Pillar 1 NHS and Pillar 2 LFD (Figures 2B and 2D).

**Figure 2:**
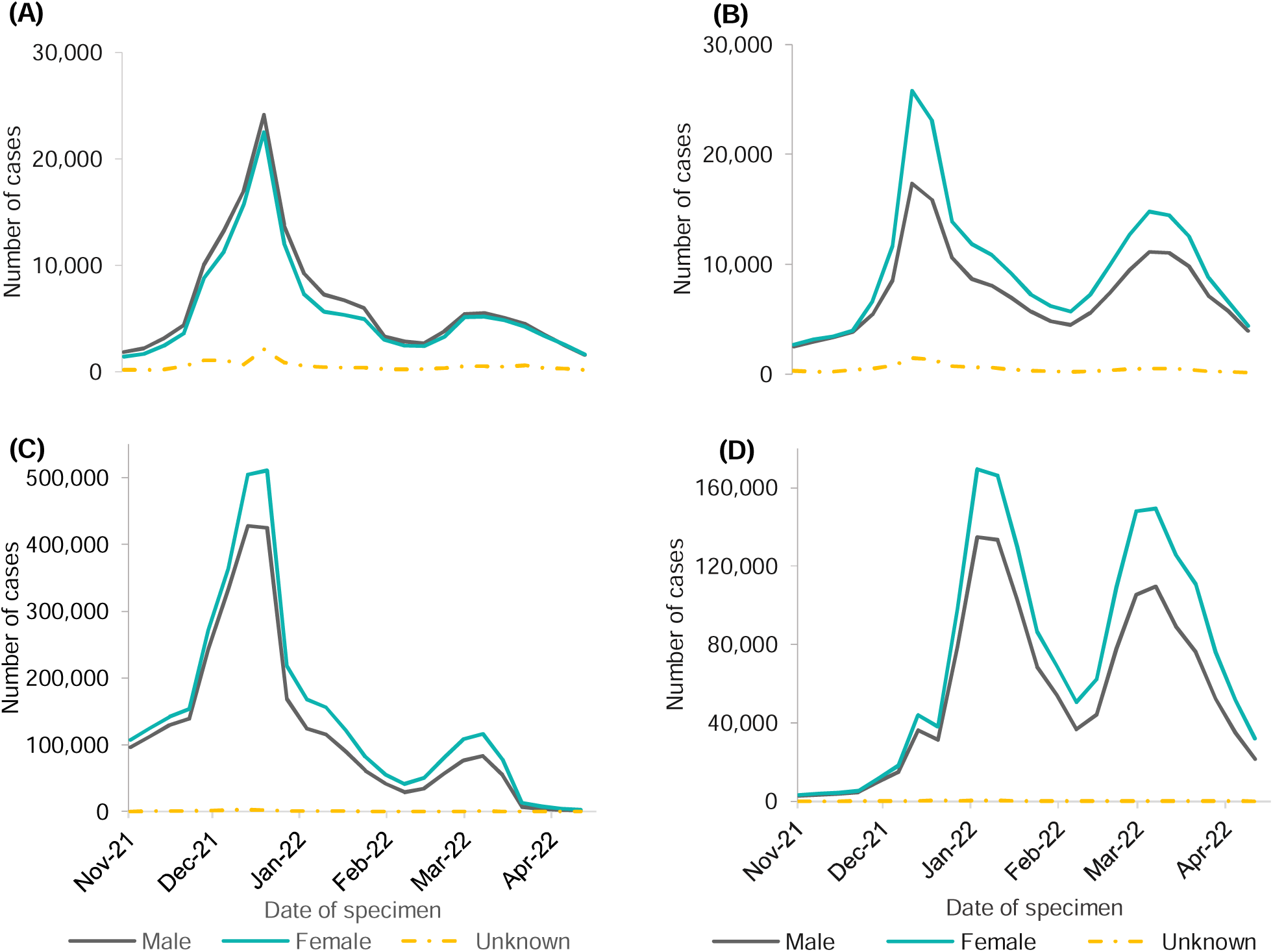
Number of weekly COVID-19 cases* tested under Pillar 1 private testing (A), Pillar 1 NHS testing (B), Pillar 2 PCR and PCR with LFD (C), Pillar 2 LFD only (D) by sex: 15 November 2021 to 30 April 2022, England. *\* Please note Figures 2A-D display crude case numbers without taking into account population denominators*.

### IMD (index of multiple deprivation; 1 – most deprived to 5 – least deprived)

The IMD trends of individuals testing under Pillar 1 NHS and Pillar 2 using LFDs appear to show opposing trends, as highlighted in Figure 3A and 3B.

**Figure 3:**
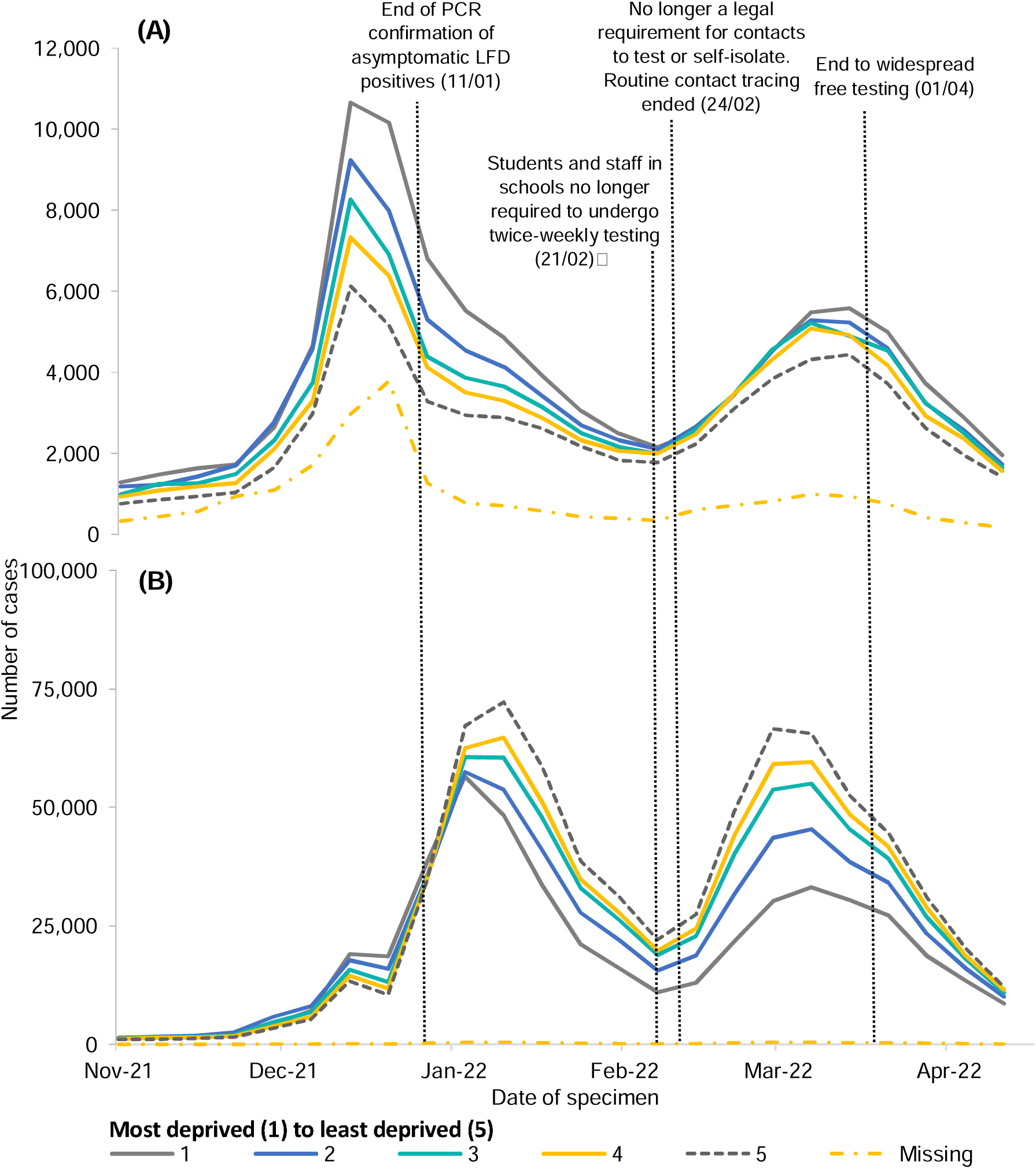
Number of weekly COVID-19 cases* tested under Pillar 1 - NHS testing (A), and Pillar 2 - LFD testing (B), by IMD quintile: 15 November 2021 to 30 April 2022, England. *Most deprived (1) to least deprived (5)* *Dates of interest due to changes in testing policy are labelled*. *\*Please note Figures 5A-B display crude case numbers without taking into account population denominators*.

The highest case numbers reported under Pillar 1 NHS testing (Figure 3A) are seen in the most deprived quintile (1^st^ IMD) from January 2022, yet for Pillar 2 LFD detected cases (Figure 3B), the highest case numbers are seen in the least deprived quintile (5^th^ IMD).

Figure 3B follows a similar trend to Figure 3A up until 11 January 2022 when confirmatory PCRs following positive LFD tests ceased. Up until this point, in both Pillar 1 NHS tested and Pillar 2 LFD tested cases, the most deprived quintiles saw the highest case numbers. After this date however, the trend reversed in those tested by LFD (Figure 3B), where the highest case numbers were seen in the least deprived and lowest case numbers in the most deprived. These observations are reinforced by age standardised COVID-19 incidence rates by quintiles of deprivation.

Figures 4B and 4D highlight an IMD disparity between those reported under Pillar 1 NHS and Pillar 2 with LFDs. The highest case rates in Figure 4B are seen in the most deprived quintile (1^st^ IMD). However, Figure 4D shows the opposite for LFD tested cases, with the highest case rates seen in the least deprived quintile (5^th^ IMD).

**Figure 4:**
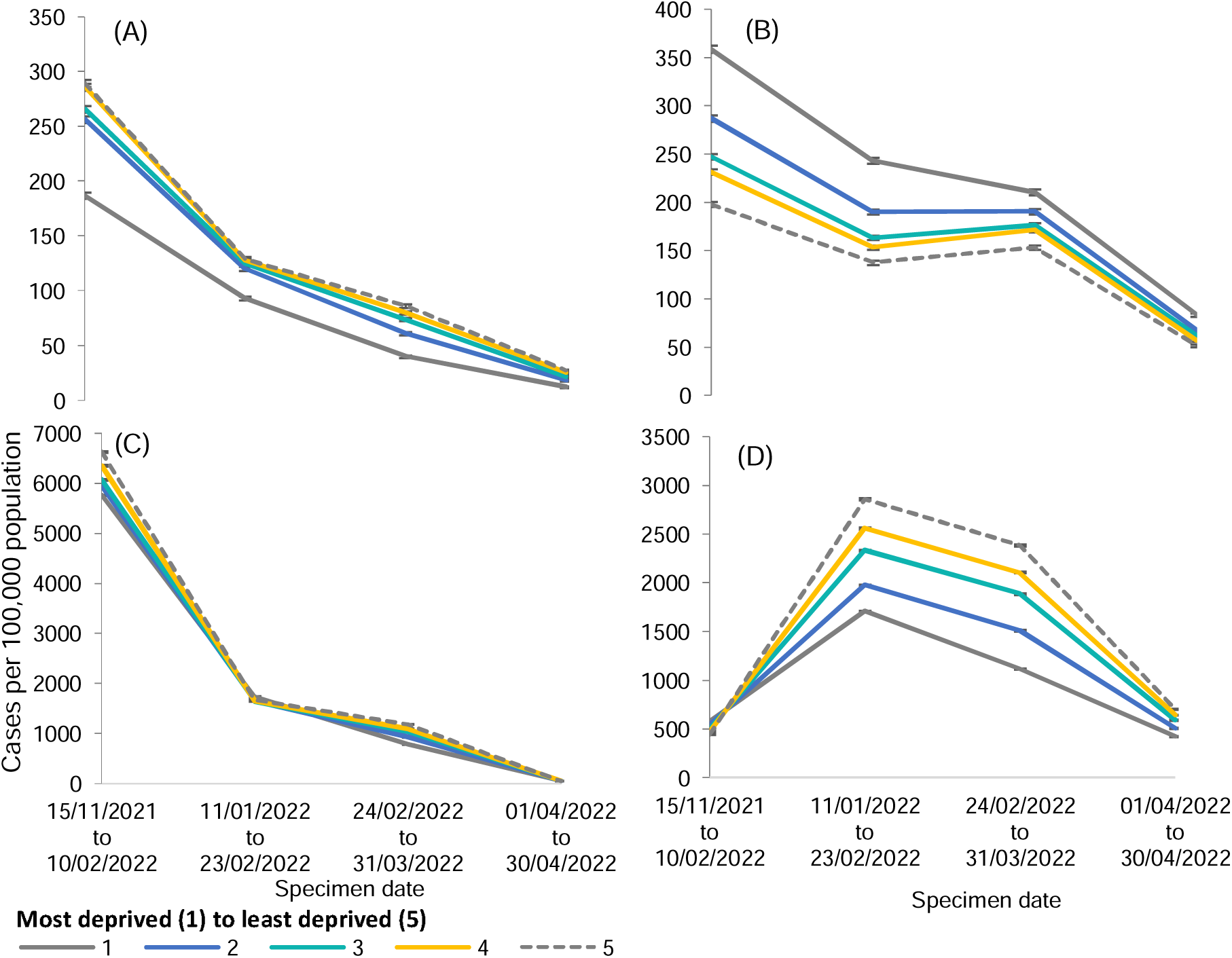
Age-standardised incidence rates of COVID-19 cases per 100,000 population tested under Pillar 1 private testing (A), Pillar 1 NHS testing (B), Pillar 2 PCR and PCR with LFDs (C), and Pillar 2 LFD tested only (D), by IMD quintile* (with 95% confidence intervals): 15 November 2021 to 30 April 2022, England. *\*Most deprived (1) to least deprived (5)*

### PHEC region

After stratifying by region, the North East had the highest age standardised incidence rates between December 2021 and 30 April 2022, except for a brief three weeks in March 2022 where the South West saw higher case rates. Over all Pillars, the highest case rate was seen in the North East region, tested under Pillar 2 by PCR and PCR with LFDs, in the first week of January 2022 (2,131 cases per 100,000 population between 3 and 9 January 2022).

Since the end to widespread free testing from the 1 to 30 April 2022, case rates in all regions tested under Pillar 2 by PCR have fallen to below 20 cases per 100,000 population. The highest case rates since 1 April are in those tested under Pillar 2 by LFD, as shown in Figure 5, however the rates are steadily declining.

**Figure 5:**
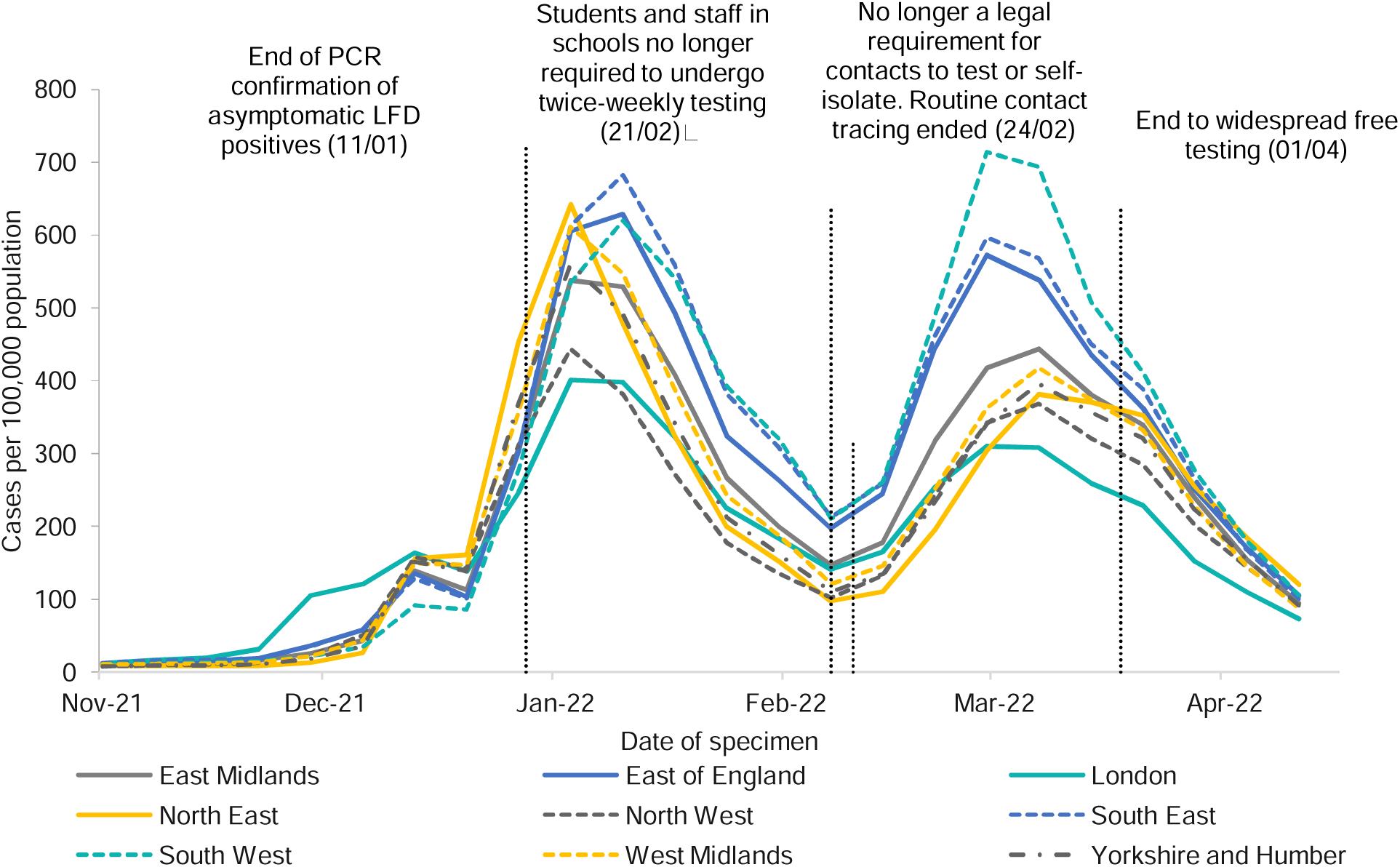
Incidence rates of COVID-19 cases per 100,000 population tested under Pillar 2 by LFD by PHEC region: 15 November 2021 to 30 April 2022, England. *Dates of interest due to changes in testing policy are labelled*.

Trends in regional incidence rates of Pillar 2 LFD detected cases show that prior to the first changes in testing in January 2022, many of the regional rates were closely overlapping. However, as further changes were introduced, the South West, South East and East of England regions diverged from other regions, with higher incidence rates of Pillar 2 LFD detected cases.

### Population density quintiles (People per square km; ranked 1 - most dense, to 5 - least dense)

In all breakdowns by pillar, those in the most densely populated areas had the highest crude case numbers, except for those tested under Pillar 2 by LFD (Figure 6), where the highest case numbers from January 2022 were seen in the least densely populated areas. This coincides with the change to testing policy ending PCR confirmatory tests after a positive LFD, on the 11 January, and just before this date the trends follow a similar pattern to other testing pillars with the most densely populated areas seeing the highest case numbers.

**Figure 6:**
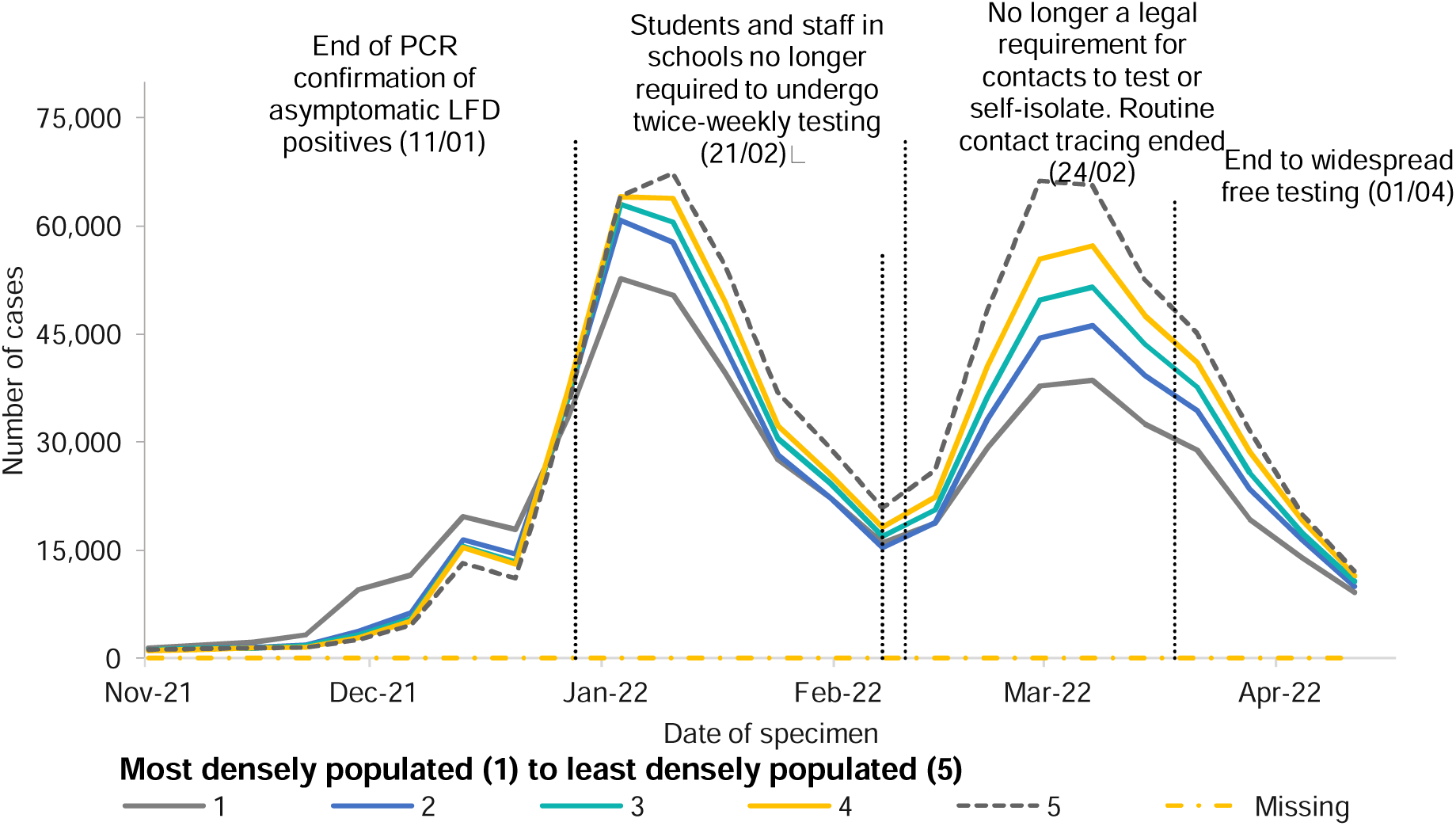
Number of Weekly COVID-19 cases tested under Pillar 2 - LFD only, by population density quintile^*^: 15 November 2021 to 30 April 2022, England. ^*\**^*people per square km; most densely populated (1) to least densely populated (5) Dates of interest due to changes in testing policy are labelled*.

## Discussion

This analysis described the demographics of reported COVID-19 cases in England, and the trends seen across different testing pillars and test types in the five months before access to free mass population testing ended. The change in Pillar 2 cases to being predominantly based on self-reporting of lateral flow tests posed new questions in how to interpret cases detected and reported through this route.

Our exploration of the distribution of cases by quintile of deprivation, both in terms of counts and age-adjusted incidence rates, indicated a divergence between Pillar 1 NHS laboratories and Pillar 2 LFDs; the former demonstrated a consistency of trends with the most deprived quintile having highest counts and rates while in contrast, Pillar 2 LFDs showed highest cases and incidence rates for the least deprived quintile.

This contrast between the two sources highlights their key differences. Pillar 1 cases are based predominantly on those tested for a clinical or public health need (such as hospitalisation or emergency department attendance) and therefore the necessity of the clinical need may overcome socio-economic factors which might affect uptake of testing outside of health services.

In addition, the continued reporting of positive results from Pillar 2 LFD tests will represent individuals or families who have specific testing behaviours and are particularly engaged with reporting their results; this is particularly notable given that free supply of LFD tests has now concluded for all but a few with specific exceptions.

It is possible that the contrast in cases by deprivation described between the two testing pillars is highlighting inequalities in the availability of information on free LFD tests, when these were available, or that test seeking behaviours may differ by the quintile of deprivation. For example, policies in supporting sick pay have varied throughout the pandemic, with COVID-19 Statutory Sick Pay and Employment and Support Allowance coming to an end on 24 March 2022, and this may have impacted behaviours in accessing COVID-19 testing after this date [3].

It will be critical to characterise reported cases from Pillar 2 in terms of their representativeness of LFD cases and infections in the community more generally. This would require the use of a range of data sources such as comparison with the ONS COVID-19 Infection Survey[11], when available. The Pillar 2 LFD data can then be contextualised and provide a valuable insight into the groups represented.

Changes in Pillar 2 LFD case distributions were also identified when examining quintiles of population density with the highest case counts transitioning from the most densely populated quintile in early December 2021 through to the least densely populated quintile, particularly following the end of regular testing recommendations for school pupils and staff.

These observations may be partially explained by the transition in the regions with the highest rates of LFD cases, with the South West, South East and East of England regions being those with the highest LFD case rates following changes to testing being introduced.

This analysis solely investigates case trends around the time policy changes to testing occurred in late 2021 and early 2022. Though testing policies are the main contributors to accessing LFD and PCR tests, there are external factors that may also have an impact that need to be assessed in other analyses. These include, evaluating unmet demand on LFD and PCR orders in periods of high prevalence such as Christmas 2021; the impact of the vaccination roll-out by age cohorts and vaccine effectiveness; and travel policy changes not only in the UK but also outside of the UK, which would have impacted the use of private testing.

Our study highlights the importance of understanding the impact of policy changes related to case detection and that such changes may impact certain population groups differently, by looking at the demographics of individuals testing positive for SARS-CoV-2. To fully understand the trends and socio-demographic differences in testing, a more in depth understanding of testing in general would benefit, in addition to an understanding of which individuals are eligible for COVID-19 treatment and therefore free testing. However, it is apparent from our findings that the cases reported through Pillar 2 LFD are overwhelmingly of working age and the least deprived.

To meet the immediate information needs, we suggest that Pillar 1 case data is preferentially used to understand the distribution of cases with a clinical or public health need for testing; although these cases will be more clinically unwell to require engagement with health services this will overcome any limitations in monitoring potential inequalities.

More widely, England is similar to other countries that are now managing reduced availability of SARS-CoV-2 testing for the general public and the emerging challenge internationally is to ensure that the data that is received is interpreted correctly, accounting for the limitations in accessibility and discussed in the context of who is accessing testing, to inform policies that address these issues regarding testing inequalities and to effectively evaluate the true impacts of the COVID-19 pandemic.

## Data Availability

No additional data available.

